# Cervical Cancer Screening Outcomes for HIV-positive Women in the Lubombo and Manzini regions of Eswatini – Prevalence and Predictors of a Positive Visual Inspection with Acetic Acid (VIA) Screen

**DOI:** 10.1101/2023.12.12.23299870

**Authors:** Rufaro Mapaona, Victor Williams, Normusa Musarapasi, Sharon Kibwana, Thokozani Maseko, Rhinos Chekenyere, Sidumo Gumbo, Phetsile Mdluli, Hugben Byarugaba, Dileepa Galagedera, Arnold Mafukidze, Alejandra de Mendoza, Prajakta Adsul, Pido Bongomin, Christopher Loffredo, Xolisile Dlamini, Deus Bazira, Sylvia Ojoo, Samson Haumba

## Abstract

This study aimed to describe the prevalence and predictors of a positive VIA (visual inspection with acetic acid) cervical cancer screening test in women living with human immunodeficiency virus (HIV). We retrospectively analysed data from women aged ≥15 who accessed VIA screening from health facilities in the Lubombo and Manzini regions of Eswatini. Sociodemographic and clinical data from October 2020 to June 2023 were extracted from the client management information system. VIA screening outcome was categorised into negative, positive, or suspicious. A logistic regression model estimated the adjusted odds ratio (AOR) of the predictors of a positive VIA screen at p<0.05 with 95% confidence intervals. Of 23,657 participants, 60.8% (n=14,397) were from the Manzini region. The mean age was 33.3 years (standard deviation 7.0), and 33% (n=7,714) were first-time screens. The prevalence of a positive VIA was 2.6% (95% CI: 2.2%, 3.0%): 2.8% (95% CI: 2.2%, 3.5%) in Lubombo and 2.4% (95% CI: 2.0%, 2.9%) in Manzini (p=0.096). Screening at mission-owned (OR 1.40; p=0.001), NGO-owned (OR 3.08; p<0.001) and industrial/workplace-owned health facilities (OR 2.37; p=0.044) were associated with positive test results compared to government-owned health facilities, and being within the 35–44 age group (OR 1.26; p=0.017) compared to 25-34 years age group was a positive predictor of a positive VIA screen. Negative predictors of positive VIA test were: being on anti-retroviral therapy (ART) for 5-9 years (OR 0.76; p=0.004) and ≥10 years (OR 0.66; p=0.002) compared to <5 years; and having an undetectable viral load (OR 0.39; p<0.001) compared to unsuppressed. Longer duration on ART and an undetectable viral load reduced the odds, while middle-aged women and screening at non-public health facilities increased the odds of a positive VIA screen.

## Introduction

Cervical cancer continues to be a leading cause of morbidity and mortality for women, despite being preventable by vaccination against the human papillomavirus (HPV) and curable if detected and treated early. There were an estimated 604,000 new cervical cancer cases and 342,000 deaths worldwide in 2020, and it is the fourth most frequently diagnosed cancer in women [1].

There are substantial inequalities in the global cervical cancer burden: incidence is three times higher in countries with lower levels of human development, and more than 90% of deaths occur in these countries. The highest regional incidence and mortality occurs in Sub-Saharan Africa, where age-standardised incidence rates in Eastern Africa (40.1), Southern Africa (36.4) and Middle Africa (31.6) [2] are far higher than the threshold of 4 per 100,000 established by the World Health Assembly’s Global Strategy for cervical cancer elimination [1].

Most cervical cancer cases (99.7%) are caused by persistent HPV infection [3], which is more prevalent in HIV-infected women [4]. The natural history of HPV infection has a slow, 10-15-year progression to pre-cancer in immuno-competent people. In HIV-infected women, the condition progresses more frequently and quickly [5]. Cervical cancer is classified as an AIDS-defining illness. It is associated with lower CD4 cell counts and a lack of anti-retroviral therapy (ART) among women living with HIV [6]. Globally, approximately 1 in 20 cervical cancers is attributable to HIV. In sub-Saharan Africa, about 1 in 5 cervical cancers is due to HIV, which threatens the gains that improving access to HIV care and treatment has made in prolonging the life expectancy of these women. However, it is essential to note that although cervical pre-cancer among women living with HIV is common, those who receive regular cervical screenings among this population have low incidence rates of invasive cervical cancer [7].

Disparities in cervical cancer incidence and mortality reflect unequal access to and coverage of comprehensive prevention (including HPV vaccination), screening and treatment of pre-cancerous lesions, and diagnosis and treatment of invasive cancers. The availability of these services is suboptimal in low- and middle-income countries. As of 2020, less than 30% of lower-middle-income countries had introduced the HPV vaccine, compared to 85% in high-income countries. Still, in 2020, less than 35% of low-income countries had national cervical cancer screening programs, and less than 30% reported the availability of pathology services, cancer surgery, and other cancer management services [8].

Secondary prevention reduces cervical cancer incidence and mortality by identifying and treating women with pre-cancerous lesions. The World Health Organisation (WHO) recommends HPV DNA testing as the primary screening test for cervical cancer, but this technology is not yet available or accessible in many countries. Cytology-based (or pap smear) screening has been successful when implemented as part of national programmes with high coverage and in settings where resources exist for patient follow-up, additional diagnostic tests (colposcopy and pathology), and disease management [9]. In low-resource settings, the "see-and-treat" approach, which includes naked eye or digitally enhanced visual inspection with acetic acid (VIA) to detect pre-cancerous lesions and the use of cryotherapy (liquid nitrous oxide ablation) to freeze and destroy pre-cancerous tissue, has been successfully implemented. However, the quality of VIA depends heavily on provider competence and the test’s sensitivity, which is variable [10–12].

### Cervical Cancer in Eswatini

Eswatini faces a high dual burden of HIV and cervical cancer. The country’s HIV prevalence of 25.9% among adults is one of the highest in the world [13], and Eswatini has the highest age-standardised cervical cancer incidence rate of 84.5 per 100,000. Only 17.6% of the country’s estimated 336,037 women aged 15 – 64 years have been screened at least once for cervical cancer [14] ^(16)^. Coverage is only slightly better for women living with HIV: estimates suggest that only 20.3% of this population has been screened [15].

Given the high dual burden of HIV and cervical cancer in the country, the Eswatini Ministry of Health has developed several policies. In particular, the National Cancer Prevention and Control Strategy of 2019 has set clear targets to increase the percentage of health facilities providing screening, early detection, and linkage to treatment for all cancers to 60% by 2022. However, only 43% of health facilities have achieved this goal. Current guidelines recommend screening as soon as one is sexually active for HIV-positive women, while HIV-negative women or those with unknown status can start screening from ≥25 years. The recommended frequency of cervical cancer screening is also based on HIV status and HPV infection status of patients (if known). Women living with HIV are screened annually, while it is every two years for HIV-negative women and extended to three years for those who are HIV and HPV-negative. All screening services are free in public hospitals. HPV DNA testing is not yet available in the country. Fidelity to these guidelines is not monitored routinely, and when done, records for non-HIV women may not be up to date. In this study, we describe the cervical cancer screening outcomes, prevalence and predictors of a positive cervical screen among HIV-positive women accessing services at selected health facilities in two regions of Eswatini.

## Methods

### Study design and setting

This was a retrospective cohort study among HIV-positive women receiving HIV care at select health facilities in the Lubombo and Manzini regions of Eswatini under the United States Government funded Support Eswatini Achieve Sustained HIV Epidemic Control (SEASEC) project. The SEASEC project is a comprehensive HIV care and treatment program implemented by the Eswatini Ministry of Health (MOH) and supported by Georgetown University. SEASEC supports the provision of cervical cancer screening and treatment of pre-cancerous lesions for all women living with HIV. As part of the program, healthcare workers have been trained to offer VIA, cryotherapy, and Loop Electrosurgical Excision Procedure (LEEP) services. Screening supplies and equipment have also been procured and installed at all seventy-five health facilities providing cervical cancer screening services in Lubombo and Manzini. Additional interventions include support for quality improvement and collection and use of service delivery data.

Health facilities in the Lubombo and Manzini regions are described in Table 1. Lubombo region has fifty-four health care facilities, including two hospitals (with in-patient services), one rural health centre, one public health unit (equivalent to a multi-department health centre), and forty-two primary health clinics. Manzini region has one hundred and twenty-three health facilities, including four hospitals (with in-patient services), two public health units (equivalent to health centres), and one hundred and seventeen primary health clinics. The SEASEC Program supports eighty-six public, private or faith-based health facilities across both regions (Table 1).

**Table 1:**
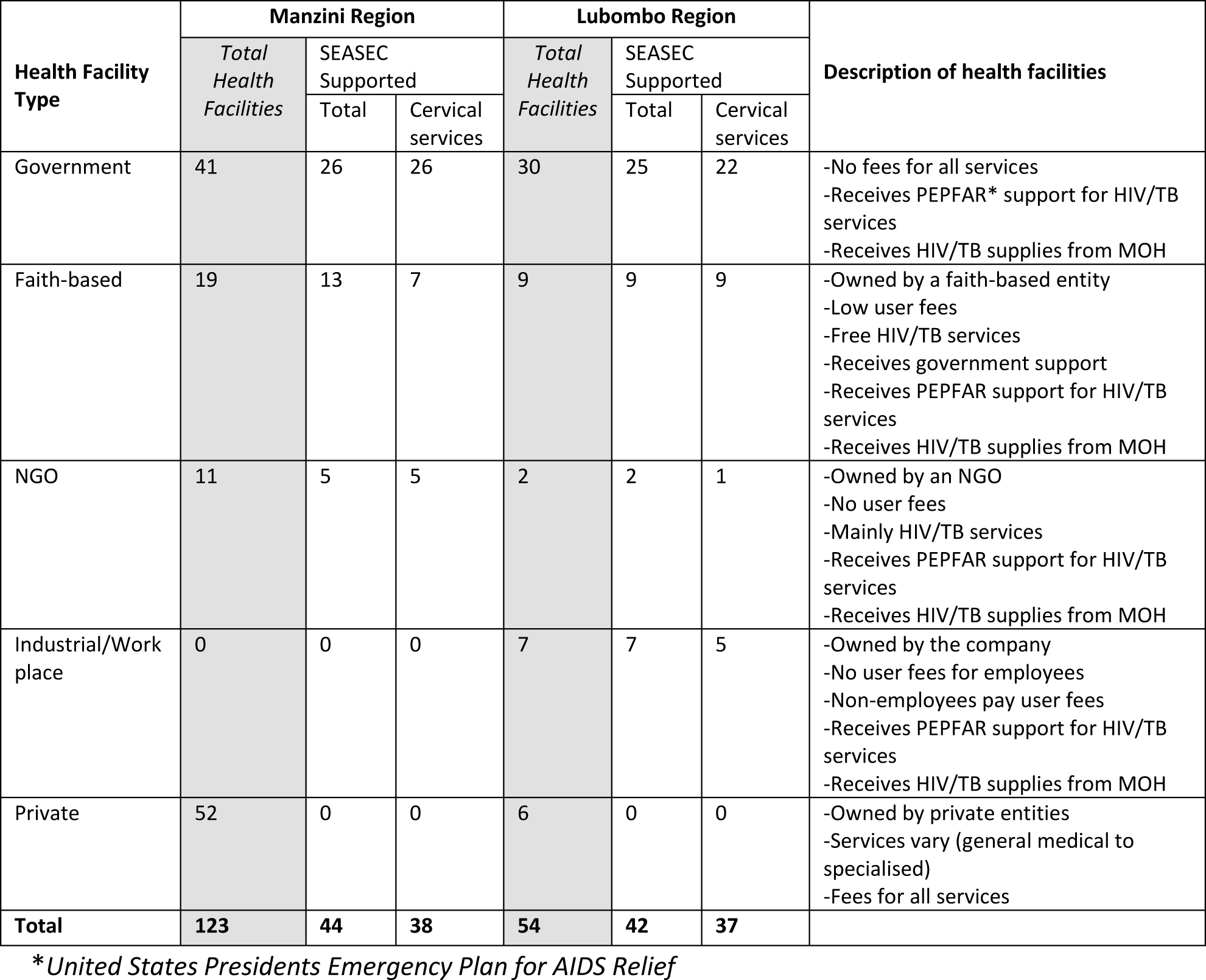
Health facilities in Manzini and Lubombo regions (Obtained from Manzini and Lubombo Regional Health Management Team Quarterly Report, Q2 2023)

| Health Facility Type | Manzini Region |  |  | Lubombo Region |  |  | Description of health facilities |
| --- | --- | --- | --- | --- | --- | --- | --- |
|  | Total Health Facilities | SEASEC Supported |  | Total Health Facilities | SEASEC Supported |  |  |
|  |  | Total | Cervical services |  | Total | Cervical services |  |
| Government | 41 | 26 | 26 | 30 | 25 | 22 | -No fees for all services<br>-Receives PEPFAR* support for HIV/TB services<br>-Receives HIV/TB supplies from MOH |
| Faith-based | 19 | 13 | 7 | 9 | 9 | 9 | -Owned by a faith-based entity<br>-Low user fees<br>-Free HIV/TB services<br>-Receives government support<br>-Receives PEPFAR support for HIV/TB services<br>-Receives HIV/TB supplies from MOH |
| NGO | 11 | 5 | 5 | 2 | 2 | 1 | -Owned by an NGO<br>-No user fees<br>-Mainly HIV/TB services<br>-Receives PEPFAR support for HIV/TB services<br>-Receives HIV/TB supplies from MOH |
| Industrial/Work place | 0 | 0 | 0 | 7 | 7 | 5 | -Owned by the company<br>-No user fees for employees<br>-Non-employees pay user fees<br>-Receives PEPFAR support for HIV/TB services<br>-Receives HIV/TB supplies from MOH |
| Private | 52 | 0 | 0 | 6 | 0 | 0 | -Owned by private entities<br>-Services vary (general medical to specialised)<br>-Fees for all services |
| Total | 123 | 44 | 38 | 54 | 42 | 37 |  |

\*United States Presidents Emergency Plan for AIDS Relief

The locations of these 75 health facilities that implement the SEASEC Program-supported cervical screening activities are shown in Figure 1.

**Figure 1:**
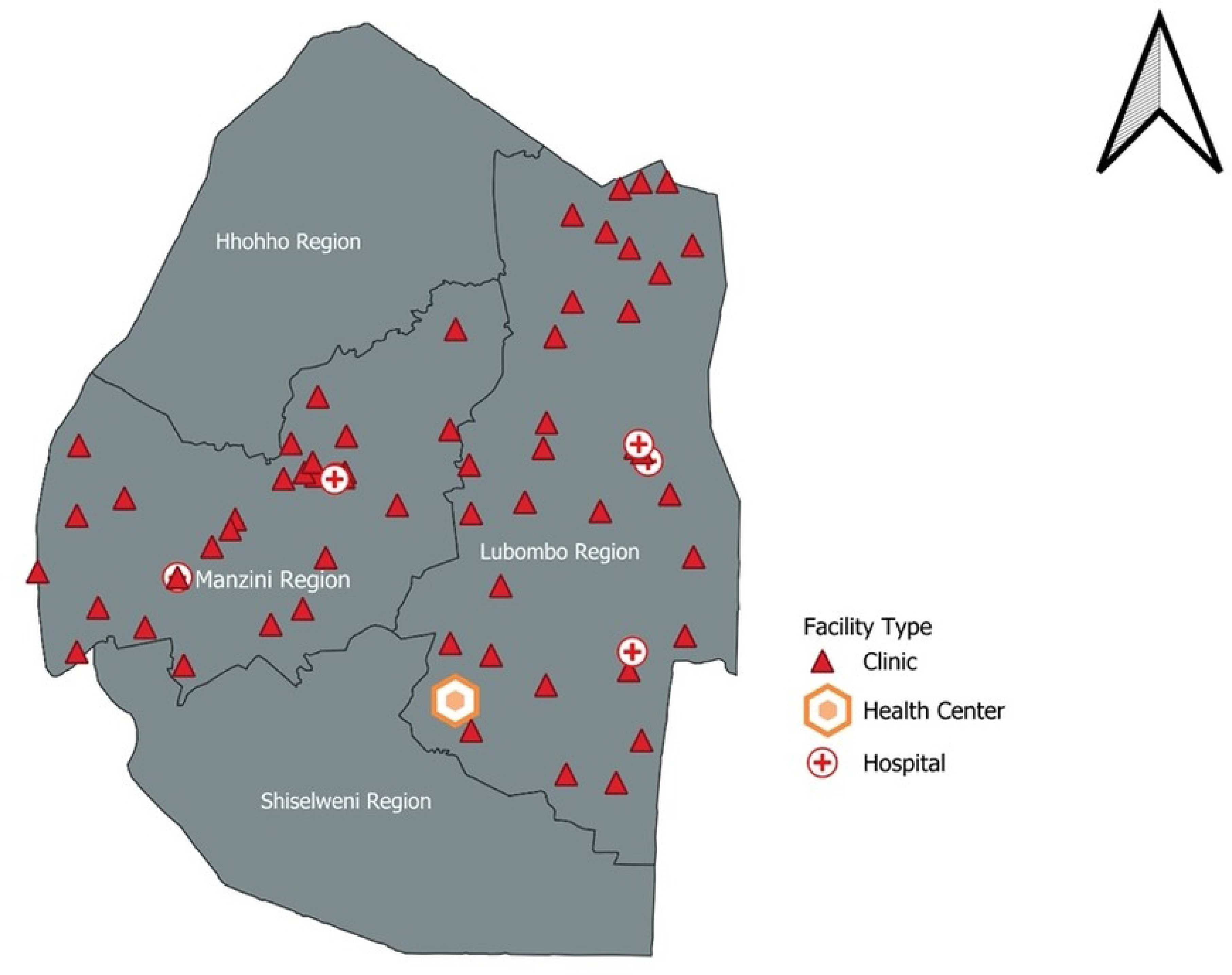
Distribution of health facilities providing cervical cancer services in the Lubombo and Manzini region supported by the SEASEC Program (Generated using SEASEC Program data)

### Study participants, sample size and sampling

We generated data for the study from patient attendance records from October 2020 to June 2023. The data are collected using standardised service registers provided by the Eswatini Ministry of Health (MOH) to all public health facilities. Our study participants were HIV-positive women screened for cervical cancer from October 2020 to June 2023 at any of the 75 health facilities described above. Data from the two regions indicate that, as of June 2023, a total of 69,529 women aged 15 years or older were living with HIV, and approximately 19,616 of these had received at least one cervical cancer screening [15]. Based on these data, a minimum of 95% of women living with HIV who have received a cervical screen (18,635) will be included in this analysis.

### Data sources and study variables

Data were extracted from the National Electronic Medical Record: Client Management Information System (CMIS) on the 2^nd^ of August 2023. The required variables were filtered and extracted per the study objectives. The variables describe the patient’s sociodemographic and clinical information (Table 2). Age was grouped into eight categories using 5-year age bands from 15 to 50 years, while parity was categorised into 0, 1, 2 – 3, 4 – 5, and ≥6. Marital status was grouped into two classes: married/living with a partner or single. The patient’s last viral load was categorised into three groups – undetectable (<50), suppressed (50<1000), and unsuppressed (≥1000). The timing of the cervical screening was divided into three groups – first-time screening, post-treatment follow-up screening, and rescreening per the national guideline. The VIA screening outcome was categorised into negative, positive, and suspicious. The outcome variable for this study is the prevalence of a positive VIA screen, defined as the proportion of women with a positive VIA screen.

**Table 2:** Descriptive characteristics of participants.

| Patient Characteristics | Negative<br>(N = 22957) | Positive<br>(N = 603) | Suspicious<br>(N = 97) | Total<br>(N = 23657) |
| --- | --- | --- | --- | --- |
| Region |  |  |  |  |
| Lubombo | 8976 (39.1%) | 256 (42.5%) | 39 (40.2%) | 9297 (39.2%) |
| Manzini | 13981 (60.9%) | 347 (57.5%) | 58 (59.8%) | 14397 (60.8%) |
| Age (years) |  |  |  |  |
| 15-19 | 284 (1.2%) | 11 (1.8%) | 0 (0.0%) | 295 (1.2%) |
| 20-24 | 2123 (9.2%) | 54 (9.0%) | 2 (2.1%) | 2194 (9.3%) |
| 25-29 | 4972 (21.7%) | 108 (17.9%) | 14 (14.4%) | 5098 (21.5%) |
| 30-34 | 5776 (25.2%) | 147 (24.4%) | 24 (24.7%) | 5949 (25.1%) |
| 35-39 | 5268 (22.9%) | 155 (25.7%) | 24 (24.7%) | 5448 (23.0%) |
| 40-44 | 3138 (13.7%) | 97 (16.1%) | 18 (18.6%) | 3268 (13.8%) |
| 45-49 | 1215 (5.3%) | 27 (4.5%) | 14 (14.4%) | 1256 (5.3%) |
| ≥50 | 181 (0.8%) | 4 (0.7%) | 1 (1.0%) | 186 (0.8%) |
| Mean (SD) | 33.29 (7.06) | 33.53 (7.06) | 36.47 (6.78) | 33.31 (7.06) |
| Parity |  |  |  |  |
| 0 | 9078 (55.9%) | 238 (57.2%) | 33 (51.6%) | 9373 (55.9%) |
| 1 | 2295 (14.1%) | 61 (14.7%) | 14 (21.9%) | 2377 (14.2%) |
| 2-3 | 3687 (22.7%) | 89 (21.4%) | 12 (18.8%) | 3789 (22.6%) |
| 4-5 | 1010 (6.2%) | 22 (5.3%) | 1 (1.6%) | 1038 (6.2%) |
| ≥6 | 179 (1.1%) | 6 (1.4%) | 4 (6.3%) | 189 (1.1%) |
| Median (Q1, Q3) | 0.0 (0.0, 2.0) | 0.0 (0.0, 2.0) | 0.0 (0.0, 2.0) | 0.0 (0.0, 2.0) |
| Duration on ART (Years) |  |  |  |  |
| Median (Q1, Q3) | 6.0 (4.0, 8.0) | 6.0 (3.0, 9.0) | 7.0 (4.0, 8.0) | 6.0 (4.0, 9.0) |
| Facility Type |  |  |  |  |
| Government | 12666 (55.2%) | 260 (43.1%) | 49 (50.5%) | 13003 (54.9%) |
| Faith-based | 6760 (29.4%) | 180 (29.9%) | 32 (33.0%) | 6981 (29.5%) |
| NGO | 3420 (14.9%) | 157 (26.0%) | 11 (11.3%) | 3588 (15.1%) |
| Industrial/Workplace | 111 (0.5%) | 6 (1.0%) | 5 (5.2%) | 122 (0.5%) |
| Cervical Screening Type |  |  |  |  |
| First Time Screening | 7427 (32.4%) | 244 (42.0%) | 29 (30.9%) | 7714 (32.6%) |
| No Outcome | 10 (0.0%) | 22 (3.8%) | 2 (2.1%) | 34 (0.1%) |
| Post Tx FU | 70 (0.3%) | 34 (5.9%) | 1 (1.1%) | 105 (0.4%) |
| Rescreening | 15432 (67.3%) | 281 (48.4%) | 62 (66.0%) | 15798 (66.8%) |
| Marital status |  |  |  |  |
| Married/living with partner | 4455 (30.5%) | 131 (34.8%) | 30 (56.6%) | 4634 (30.7%) |
| Single | 10164 (69.5%) | 245 (65.2%) | 23 (43.4%) | 10451 (69.3%) |
| Last VL Outcome (cells/ml) |  |  |  |  |
| No VL Result | 657 (2.9%) | 12 (2.0%) | 2 (2.1%) | 671 (2.8%) |
| Suppressed (50<1000) | 2389 (10.4%) | 97 (16.1%) | 8 (8.2%) | 2494 (10.5%) |
| Unsuppressed (≥1000) | 406 (1.8%) | 25 (4.1%) | 3 (3.1%) | 434 (1.8%) |
| Undetectable (<50) | 19505 (85.0%) | 469 (77.8%) | 84 (86.6%) | 20095 (84.8%) |

### Statistical analysis

Data was extracted in .xls format and imported into Stata 17 (College Station, TX) for cleaning and analysis. Descriptive analysis summarised underlying trends and seasonal patterns. Key patient characteristics are presented in Table 2, disaggregated by the cervical screening outcome. Casewise analysis was used since all the patients had the outcome of interest. The prevalence of positive VIA screens was presented overall and disaggregated by other patients’ sociodemographic and clinical information.

A new binary dependent variable – presence or absence of cervical lesion - was generated to determine the predictors of a positive cervical screen. A logistic regression model was fitted using the dependent variable and the different sociodemographic and clinical variables as predictors. A univariate model was initially built using each of the independent variables, and a stepwise forward and backward elimination process was used to select variables for inclusion in the final multivariate model at p=0.20. The final predictors were determined from the multivariate model at p<0.05.

### Ethical review and approval

This study is covered under the protocol approved by the Eswatini Health and Human Research Review Board (EHHRRB 116/2022) for Georgetown University to analyse program data for dissemination. Additional approval has been obtained from the Georgetown University Institutional Review Board (GU - IRB) (STUDY 00006034) and the United States Centers for Disease Control (CDC) (Accession #: CGH-ESW-9/14/23-15af6). Anonymised data were used to ensure confidentiality and the EHHRRB also approved an application for a waiver of written informed consent from participants since the data is from routine care extracted from the electronic medical records.

## Results

### Sociodemographic and screening outcome characteristics of study participants

Table 2 presents the descriptive characteristics of all HIV-positive patients included in the study and disaggregated by screening outcomes (negative, positive, and suspicious cervical screens). Overall, 23,657 patients accessed cervical screening services. Six hundred and three (2.6%) had a positive screen, and 60.8% (n=14, 397) were from the Manzini region. The mean age was 33.3 years (SD 7.0). Analysis by 5-year categories showed that screening increased gradually from 1.2% (n= 295) in the 15 – 19 age group to 25% (n=5949) in the 30 – 34 age group and was consistent for both positive and negative participants. Most patients (67.3%, n=15,798) were rescreened, while 32.6% (n=7,714) were screened for the first time. About 69% (n=10, 451) were single, and more than half of the patients (55.9%, n=9,373) had a parity of 0. This was similar across the three outcome groups. The majority (96.4%, n=22,770) of the patients were on a dolutegravir-based ART regimen; the median duration of ART was 6.0 years (IQR 4.0, 9.0), and 95% overall had a suppressed and undetectable viral load. This is similar for those who screened positive (94%, n=566).

### Prevalence of positive VIA screen

The overall prevalence of a positive cervical screen was 2.6% (95% CI: 2.2%, 3.0%): 2.8% (95% CI: 2.2%, 3.5%) in Lubombo and 2.4% (95% CI: 2.0%, 2.9%) in Manzini (p=0.096). The prevalence was further analysed by age group, ART duration, parity, and cervical screening type (Table 3). The prevalence ranged from 2.1% (95% CI: 1.5%, 3.0%) in the 25 – 29 years age group to 3.7% (95% CI: 1.3%, 9.9%) in the 15 - 19 years age group, and it was statistically higher in the 40 - 44 years age group in Lubombo (4.2%; 95% CI: 2.5%, 6.8%) compared to those in Manzini (2.1%; 95% CI: 1.4%, 3.1%) (p=0.001). Patients with parity ≥6 had an overall higher prevalence of 3.2% (95% CI: 0.5%, 19.5%), while those with parity=0 had a higher prevalence in Lubombo compared to Manzini (p=0.004). Prevalence was higher in patients receiving post-treatment follow-up screening at 32.7% (95% CI: 20%, 49%) and lowest in rescreening patients.

**Table 3:** Prevalence of a positive cervical screen.

| Patient Characteristics | % Prevalence (95% Confidence Interval) |  |  | P-Value* |
| --- | --- | --- | --- | --- |
|  | Lubombo | Manzini | Overall |  |
| Region | 2.8% [2.2,3.5] | 2.4% [2.0,2.9] | 2.6% [2.2,3.0] |  |
| Facility Type |  |  |  |  |
| Government | 2.1% [1.6,2.8] | 1.9% [1.4,2.6] | 2.0% [1.6,2.5] | **0.440** |
| Mission | 4.7% [3.2,7.0] | 1.6% [1.0,2.6] | 2.6% [1.9,3.5] | **<0.001** |
| NGO | 0.0% | 4.4% [3.5,5.6] | 4.4% [3.5,5.6] | - |
| Industrial/Workplace | 5.1% [2.3,10.9] | 0.0% | 5.1% [2.3,10.9] | - |
| Age group (years) |  |  |  |  |
| 15-19 | 5.9% [1.4,21.7] | 2.3% [0.7,7.4] | 3.7% [1.3,9.9] | 0.108 |
| 20-24 | 2.4% [1.1,5.2] | 2.6% [1.5,4.4] | 2.5% [1.6,3.9] | 0.789 |
| 25-29 | 2.4% [1.3,4.2] | 2.0% [1.2,3.2] | 2.1% [1.5,3.0] | 0.389 |
| 30-34 | 2.6% [1.5,4.4] | 2.4% [1.6,3.6] | 2.5% [1.8,3.4] | 0.632 |
| 35-39 | 2.4% [1.4,4.1] | 3.1% [2.3,4.3] | 2.9% [2.2,3.8] | 0.136 |
| 40-44 | 4.2% [2.5,6.8] | 2.1% [1.4,3.1] | 3.0% [2.1,4.2] | **0.001** |
| 45-49 | 2.1% [1.0,4.7] | 2.2% [1.3,3.6] | 2.2% [1.4,3.4] | 0.932 |
| ≥50 | 4.9% [1.8,12.3] | 0.0% | 2.2% [0.8,5.6] | - |
| ART duration |  |  |  |  |
| <5 years | 3.2% [2.2,4.8] | 2.8% [2.2,3.7] | 3% [2.4,3.7] | 0.332 |
| 5-9years | 2.5% [1.7,3.6] | 2.3% [1.7,3.1] | 2.4% [1.9,3.0] | 0.425 |
| ≥10years | 2.8% [1.8,4.4] | 2.0% [1.4,2.8] | 2.3% [1.7,3.2] | 0.070 |
| Parity |  |  |  |  |
| 0 | 3.2% [2.2,4.6] | 2.2% [1.6,3.0] | 2.6% [2.0,3.2] | **0.004** |
| 1 | 3.1% [1.6,5.9] | 2.2% [1.1,4.4] | 2.6% [1.6,4.1] | 0.184 |
| 2-3 | 2.7% [1.4,5.0] | 2.1% [1.2,3.6] | 2.4% [1.6,3.5] | 0.272 |
| 4-5 | 2.0% [0.8,5.0] | 2.3% [0.7,7.0] | 2.1% [1.0,4.4] | 0.768 |
| ≥6 | 4.7% [0.7,26.4] | 0.0% | 3.2% [0.5,19.5] | 0.097 |
| Frequency of screening |  |  |  |  |
| First Time Screening | 4.0% [2.4,6.5] | 2.9% [2.2,3.9] | 3.2% [2.5,4.1] | 0.068 |
| Post Treatment Follow-Up | 37.1% [16.5,64] | 30.4% [15.6,50.9] | 32.7% [20,49.0] | 0.491 |
| Rescreening | 1.9% [1.4,2.6] | 1.7% [1.3,2.1] | 1.8% [1.5,2.2] | 0.235 |

\*Compares Lubombo and Manzini

### Follow-up VIA screening

Two hundred and twenty-three of 603 patients with a positive first VIA result had follow-up VIA results. Of the 223, 125 (56%) had a negative result, 89 (40%) had a positive result, and 9 (4%) had a suspicious result. Only 21 of 97 patients with a suspicious result had a follow-up VIA result: 5 (23.8%) were negative, 3 (14.3%) were positive, and 13 (61.9%) remained suspicious. Of 22,957 patients with a negative VIA result, follow-up screening data was available for 8403 patients: 8,288 (98.6%) remained negative, 50 (0.6%) were positive, and 13 (0.15%) had a suspicious result.

### Predictors of a positive cervical screen

The predictors of a positive cervical screen are presented in Table 4. In the univariate analysis, screenings at mission-owned, NGO-owned, and industrial/workplace-owned health facilities were significant positive predictors of a positive cervical cancer screening result compared to screening at a government-owned health facility. Being on ART for 5 – 9 years and ≥10 years compared to being on ART for less than 5 years and having an undetectable viral load compared to an unsuppressed viral load were significant negative predictors of a positive cervical screen.

**Table 4:**
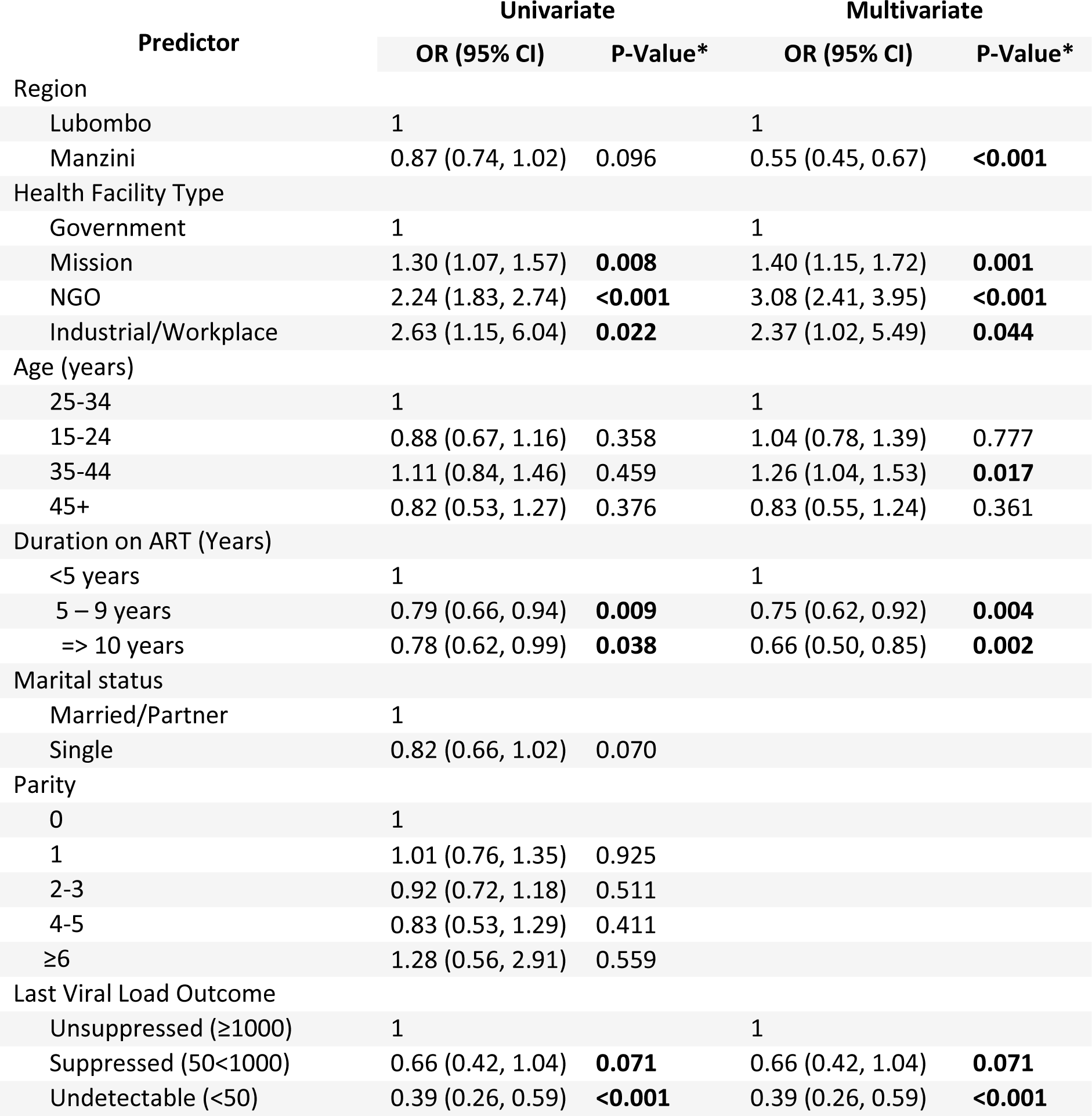
Univariate and Multivariate Predictors of Abnormal Cervical Screen.

In the multivariate analysis, significant negative predictors of a positive cervical screen were residing in the Manzini region (OR 0.55; 95% CI: 0.45, 0.67; p<0.001) compared to Lubombo; being on ART for 5 – 9 years duration (OR 0.76; 95% CI: 0.62, 0.92; p=0.004) and ≥10 years (OR 0.66; 95% CI: 0.50, 0.85; p=0.002) compared to less than 5 years; and having an undetectable viral load result (OR 0.39; 95% CI: 0.26, 0.59; p<0.001) compared to an unsuppressed viral load. Being screened at mission-owned (OR 1.40; 95% CI: 1.15, 1.72; p=0.001), NGO-owned (OR 3.08; 95% CI: 2.41, 3.95; p<0.001) and industrial/workplace-owned (OR 2.37; 95% CI: 1.02, 5.49; p=0.044) compared to government-owned health facilities, and being within the 35 – 44 age group (OR 1.26; 95% CI: 1.04, 1.53; p=0.017) compared to 25-34 years age group, were significant positive predictors of a positive cervical screen.

## Discussion

We aimed to describe the cervical cancer screening outcomes, prevalence, and predictors of a positive cervical screen in the women living with HIV who accessed services at select health facilities in the Lubombo and Manzini regions of Eswatini. Our study findings suggest that the percent positivity for cervical cancer screening tests among WLHIV is 2.6%, lower than reported in the literature from studies in similar settings, regardless of the type of screening test used. A previous cross-sectional study in Eswatini found that the presence of cervical lesions was higher in HIV-positive (22.9%) than HIV-negative women (5.7%; p < 0.0001) [16], and another study in Eswatini found a VIA positivity rate of 9% in the general population [17]. Elsewhere in Africa, a large cohort study of women in Zambia who were screened with VIA and digital cervicography (VIAC) had a positivity rate of 10.4% for the general population, with WLHIV having a much higher rate of 53.3% [18]. In Nigeria, a study using program data found a VIA positivity rate of 7.1% [19]. In another study assessing HPV testing for WLHIV, 46.5% in Burkina Faso and 43.8% in South Africa had a positive HPV test [20]. In a hospital-based study conducted in Ethiopia, the prevalence of a pre-cancerous cervical lesion was 9.3% among WLHIV [21].

This finding of a lower-than-expected positivity rate in Eswatini compared to the studies cited above points to the need for further research into the screening offered in health facilities. Given the literature around VIA screening, the learning curve to implement this screening test is high, and the results during initial phases often reflect lower positivity rates. Future research can help determine whether the lower-than-expected positivity rate is due to a need for strengthening screening services or is an accurate reflection of rates among this population. Given the move towards the recent WHO guidelines in Eswatini, HPV-DNA is being recommended for use as the primary screening test. Eswatini has yet to fully roll out this testing modality; hence, ensuring the quality of the existing VIA screening test is essential even as the government works to expand access to HPV DNA testing.

Another observation from our study is that the prevalence of a positive screening result was associated with accessing services at faith-based, NGO-owned and industrial/workplace-owned health facilities. This observation could be due to several reasons. First, personnel at these types of health facilities who provide cervical screening services are assigned to VIA screening as part of their ongoing, long-term responsibilities, which enables them to acquire skills and expertise over time. In comparison, health personnel at government-owned health facilities are not stationed at the VIA unit; they rotate from their role to another unit or health facility every twelve months. This rotation limits the skill and expertise in cervical screening. Secondly, the facilities with more positive screens have access to better screening equipment with better utilisation than government-owned facilities. Third, these health facilities enforce higher quality standards on VIA screening with strict adherence to the screening standard operating procedure (SOP) and supervision than government-owned health facilities. Fourth, they provide supplemental training for their staff in addition to the required minimum training for health personnel providing VIA and cervical cancer services.

A positive screening result in our study was also associated with being aged between 35 – 44 years, compared to older and younger age groups. This finding is consistent with findings from several sub-Saharan African countries that the burden of cervical cancer attributable to HIV is highest among younger women aged less than 45 [22]. Longer duration on ART and an undetectable viral load were protective in our analysis, which is consistent with other studies and could be attributed to the increased immune function, which reduces the incidence and progression of squamous intraepithelial lesion (SIL) and cervical intraepithelial neoplasia (CIN) and, ultimately, the incidence of invasive cervical cancer when patients are stable on ART [23]). Other predictors of positive screening results vary in the literature. Studies have found that predictors of pre-cancerous cervical lesions or positive cervical cancer screening results among WLHIV include increasing parity [24], a history of multiple sexual partners, and sexually transmitted infections [25]. In the general population, predictors of VIA positivity include having multiple sexual partners, having an early sexual debut, and being older [17,26].

Our study had several strengths, notably the large sample size of nearly 24,000 women. Regionwide coverage of public, faith-based, NGO-owned and industrial/workplace-owned health facilities limits any possible effects of clinic-based selection bias. There was a high rate of data completion on the results of screening tests, with very few participants lost to follow-up or with missing data. Given our study’s large number of health facilities, the findings highly represent the two regions. On the contrary, the study conclusions were limited in generalizability, as the SEASEC project supported all sites included in this study. These findings may not apply to settings not supported by the SEASEC project. This is also important for planning future research to outline the role of the support partners, the characteristics of the settings, and the support received. Another limitation of our study is excluding private health facilities, which are not SEASEC-supported. A challenge with private health facilities includes difficulty accessing their data, uncertainty about the type of services they offer, and whether they adhere to standard MOH guidelines.

Eswatini has yet to achieve its target of scaling up cervical cancer services. Scaling up cervical cancer services to more health facilities with targeted, demand-driven activities and patient education will increase cervical screening coverage in Eswatini. More positive screens can be identified, and women can be offered treatment with a higher coverage.

Government-owned health facilities should adopt practices to increase accountability for cervical screening services with measures to track adherence to cervical screening standards, standard operating procedures, supportive supervision, and mentoring for staff who require additional skills.

Early HIV case finding and scaling up ART uptake for women in different at-risk populations can limit non-suppression and lower the risk of a positive cervical screen. Moreover, scaling up HIV services overall will increase cervical screening opportunities.

Although Eswatini has just commenced HPV vaccinations for eligible females, the implementation has not been country-wide due to the limited availability of the vaccine. This should be scaled up to all regions with provisions to make it accessible for eligible residents of remote, hard-to-reach locations. The general public should receive adequate information on the vaccine’s benefits, including details of where and how they can access additional information on the HPV vaccines.

Finally, additional studies are required to identify other possible causes of the lower-than-expected VIA positivity rate in selected facilities with strategies to address the identified gaps. Ongoing screening with VIA should be standardised across all health facilities, focusing on improving the health system structures required for a comprehensive screening program. Efforts to expand access to HPV DNA testing should be fast-tracked, prioritising screening for vulnerable women, including WLHIV.

## Data Availability

The data underlying this research is not readily available for sharing at the moment but will be made available soon as relevant approvals are obtained.

## Acknowledgements

We thank all the clients and healthcare workers, without which this study would not have been possible. We also thank the monitoring and evaluation team at Georgetown University Eswatini, who diligently collected the data reported in the article.

## Contributions

RM, VW, SO and SH conceptualised the study. NM, TM, RC, SG, PM, HB, and DG participated in data capturing and verification at health facilities supervised by VW and AM. VW, AM and TM conducted the data analysis. RM, VW, and SG wrote the first draft of the manuscript. SK, ADM, PA, PB, CL, XD, DB, and SO contributed substantially to manuscript revisions. All authors contributed to interpreting the results and reviewing different drafts of the manuscript. All authors read and approved the final draft.

## Declarations

### Data availability

The data presented in this research are available upon a reasonable request to the corresponding author.

### Ethical review and approval

This study is covered under the protocol approved by the Eswatini Health and Human Research Review Board (EHHRRB 116/2022) for Georgetown University to analyse program data for dissemination. The EHHRRB also approved a waiver of informed consent by patients. Additional approval has been obtained from the Georgetown University Institutional Review Board (GU - IRB) (STUDY 00006034) and the United States Centers for Disease Control (CDC) (Accession #: CGH-ESW-9/14/23-15af6). Anonymised data were used to ensure confidentiality and the EHHRRB also approved an application for a waiver of written informed consent from participants since the data is from routine care extracted from the electronic medical records.

